# Estimating SARS-CoV-2 variant strain infectiousness in Japan as of March 28, 2021

**DOI:** 10.1101/2021.04.11.21255283

**Authors:** Junko Kurita, Tamie Sugawara, Yasushi Ohkusa

## Abstract

**Background:** A variant strain of SARS-CoV-2, VOC202012/01, emerged in the UK in September, 2020. Its infectiousness was estimated as higher than that of the original strain.

**Object:** We estimated the infectiousness of the variant strain of SARS-CoV-2 in comparison to that of the original strain under conditions prevailing in Japan.

**Methods:** We estimated infectiousness through a simple susceptible–infected–recovered (SIR) model by strain. The study period was March 1–28, 2020. The information used for the study was available as of April 3, 2020.

**Results:** The estimated reproduction number of the SARS-CoV-2 variant strain was 1.799; its 95% CI was [1.642, 1.938]. The onset date of the first case in Japan was estimated as December 4 ([November 16, December 14]), 2020. However, infectiousness of the original strain was estimated as 1.123 ([1.093, 1.166]).

**Discussion and Conclusion:** We demonstrated that infectiousness increased by 0.684 or 60%, increasing from the original to the variant. That finding might be comparable to that of a study conducted in the UK. However, the difference must be monitored continuously and carefully.

## Introduction

A variant strain of SARS-COV-2, VOC202012/01, emerged in the UK in September 2020. Its infectiousness and pathogenicity were estimated as 75% higher than those of the original strain, which was previously circulating before emerging variant strain[1].

In Japan, the first case was detected on January 6 as an imported case from the UK. Subsequently until the end of March, 2021, community outbreak of the variant strain occurred in Japan. Since the beginning of March, 2021, the outbreak was remarkable in Japan. Actually, the Ministry of Health, Labour and Welfare reported prevalence of the variant strain in the community outbreak as 7, 10, 16, and 20% for each week of March in Japan [2].

However, the estimated infectiousness of the variant strain was not evaluated outside Europe. Even for the outbreak caused by the original strain, outbreak situations differed among countries. For example, prevalence of SARS-COV-2, with mainly the original strain, in the USA were 9361 and 6558 cases in the UK per 100 thousand residents, but in Japan it was limited to 372 as of the end of March, 2021 [3]. Infectiousness of the variant strain might differ country-by-country, as that of the original strain has. Therefore, we evaluated the infectiousness of the variant strain compared to that of the original strain under the same conditions using a simple deterministic SIR model in Japan.

## Methods

We defined the variant strains as mutated at N501Y and/or E484K together because the numbers of patients were not reported by subtype among variant strains. Therefore, we cannot specifically examine differences among variant strains.

For a simple deterministic SIR model [4–6] applied to the epidemic curve for Japan’s 120 million populations, we used the numbers of symptomatic patients reported by strain from MHLW for March 1–28, 2021, which were published [7] on April 4, 2021. We excluded patients who were presumed to be infected abroad because those patients presumably do not represent cases of community-acquired infection in Japan. We assumed that the two strains had similar properties, except for infectiousness.

Because the SIR model is a system reflecting daily differences, we converted weekly data simply to daily data divided by 7.

The incubation period is assumed to conform to the empirical distribution in Japan.Letting the probability of onset on day *k* after exposure in the empirical distribution be *p*(*k*), then *p*(*k*)/Π_*m*=0_^*k*-1^ (1-*p*(*m*)) of infected patients showed no symptom until *k*-1 days from exposure showed onset on day *k*. That is, 1-*p*(*k*)/Π_*m*=0_^*k*-1^(1-*p*(*m*)) of them showed onset on *k*+1 day after exposure, or later.

For symptomatic patients with unknown onset date, we estimated the onset date as follows: Letting *f*(*k*) represent the probability of a *k* day delay in the empirical distribution of reporting delay from onset and letting *N*_*t*_ denote the number of patients for whom onset dates were not available for publication at date *t*, then the number of patients for whom the onset date was known is *t*−1. The number of patients for whom onset dates were not available was estimated as *f*(1)*N*_*t*_. Similarly, the number of patients with onset date *t*−2 and for whom onset dates were not available were estimated as *f*(2)*N*_*t*_. Therefore, the total number of patients for whom the onset date was not available, given an onset date of *s*, was estimated as Σ_*k*=1_*f*(*k*)*N*_*s*+*k*_ for the long duration extending from *s*.

Moreover, the reporting delay from the Ministry of Health, Labour and Welfare (MHLW) might be considerable. In other words, if *s*+*k* is larger than that in the current period *t*, then *s*+*k* represents the future for period *t*. For that reason, *N*_*s*+*k*_ is not observable. Such a reporting delay engenders underestimation bias of the number of patients. For that reason, it must be adjusted as Σ_*k=1*_^*t-s*^*f(k)N*_*s+k*_ */*Σ _*k=1*_^*t-s*^*f*(*k*). Similarly, patients for whom the onset dates were available are expected to be affected by the reporting delay. Therefore, *M*_*s*_|_*t*_ /Σ_*k=1*_^*t-s*^*f*(*k*), where *M*_*s*_|_*t*_ represents the reported number of patients for whom onset dates were within period *s*, extended until the current period *t*. We used these arranged data to produce the epidemic curve for SIR modelling.

The distribution of infectiousness in symptomatic cases was assumed to be 30% on the onset day, 20% on the following day, and 10% for the subsequent 5 days [8]. If designating the distribution of infectiousness as *g*(*k*), where *k* represents days from onset and the number of symptomatic patients on day *t* and onset was *k* days ago as *I*(*t,s*), then the number of newly infected patients on day *t* was *R*(*t*) *S*(*t*)Σ_*k=0*_ *g*(*k*) *I*(*t,k*)/*Y*(*t*), where *S*(*t*) denotes the number of susceptible population and *Y*(*t*) represents the total population on day *t*.

We run the simulation from the initial case under the presumed reproduction number. Simulation results of the epidemic curve, which was defined as the number of patients by onset date, was converted to the predicted reporting number of laboratory-confirmed patients by the empirical distribution of reporting delay from onset. To that was added a constant proportion of asymptomatic patients, 20%, assuming no infectiousness among asymptomatic patients [8].

If the number of the estimated reported laboratory-confirmed patients on a day in the simulation was higher than the actual data on March 1, then we compared the model prediction and observation for four weeks to seek reproduction number by types to achieve the minimum absolute difference among them through grid search on [1,2] by 0.0001. The bootstrapping procedure we used was fully replicated bootstrapping for a fixed number of initial cases. There were *N* patients in the data, with numbering of the patients from the initial case to the last case. Initially, no patient was on the bootstrapped epidemic curve. If a random variable drawn from a uniform distribution of (0,1) was *t* included in the internal [*i*/(*N*-1), (*i+*1)/(*N*-1)], then we added one to the onset date of the *i*+1th patient to the bootstrapped epidemic curve. We replicated this procedure *N*-1 times. Thereby, we obtain the bootstrapped epidemic curve with *N*-1 patients. Finally, we added the initial patient, for whom the onset date was January 14, to the bootstrapped epidemic curve. Consequently, we obtained a bootstrapped epidemic curve with *N* patients.

The study period was March 1–28, 2021 for estimating the infectiousness of the two strains. The information used for the study was available on April 4, 2021. The empirical distribution of the incubation period and reporting delay were based on the outbreak in Japan from January 14, 2020 to the end of February, 2021.

## Results

In the four weeks I March 2021, 7264, 7940, 8960 and 12045 laboratory-confirmed cases including variant strain cases as well as the original strain cases were reported [7]. Multiplying proportion of the variant strain described above to these number, the weekly number of reported laboratory-confirmed patients by strain in March, 2021 were shown in Figure 1. One seventh of these data were used for the following analysis as daily data.

**Fig. 1.**
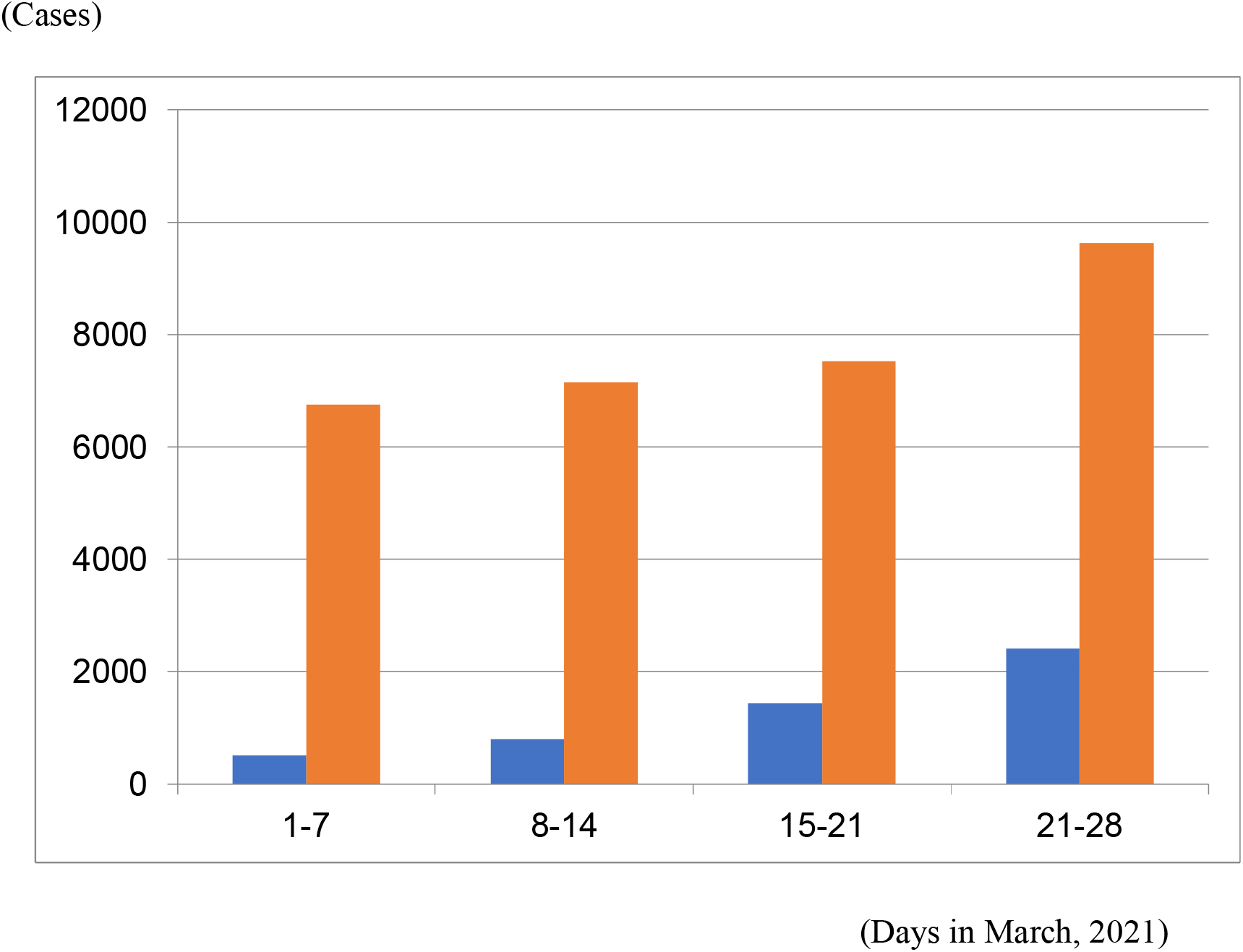
Number of laboratory confirmed cases infected by original and variant strain of SARS-CoV-2 in March, 2021 reported by MHLW, Japan. Note: Orange bars represent numbers of laboratory-confirmed cases infected by the original strain weekly. Blue bars represent patients infected by the variant strain. Laboratory-confirmed cases included asymptomatic and presymptomatic cases. Dates are the reported date: not the onset or infected date.

Figure 2 presents an empirical distribution of the duration of onset to reporting in Japan. The maximum delay was 31 days. Figure 3 depicts the empirical distribution of incubation periods among 125 cases for which the exposed date and onset date were published by MHLW in Japan. The mode was 6 days. The average was 6.6 days.

**Fig. 2.**
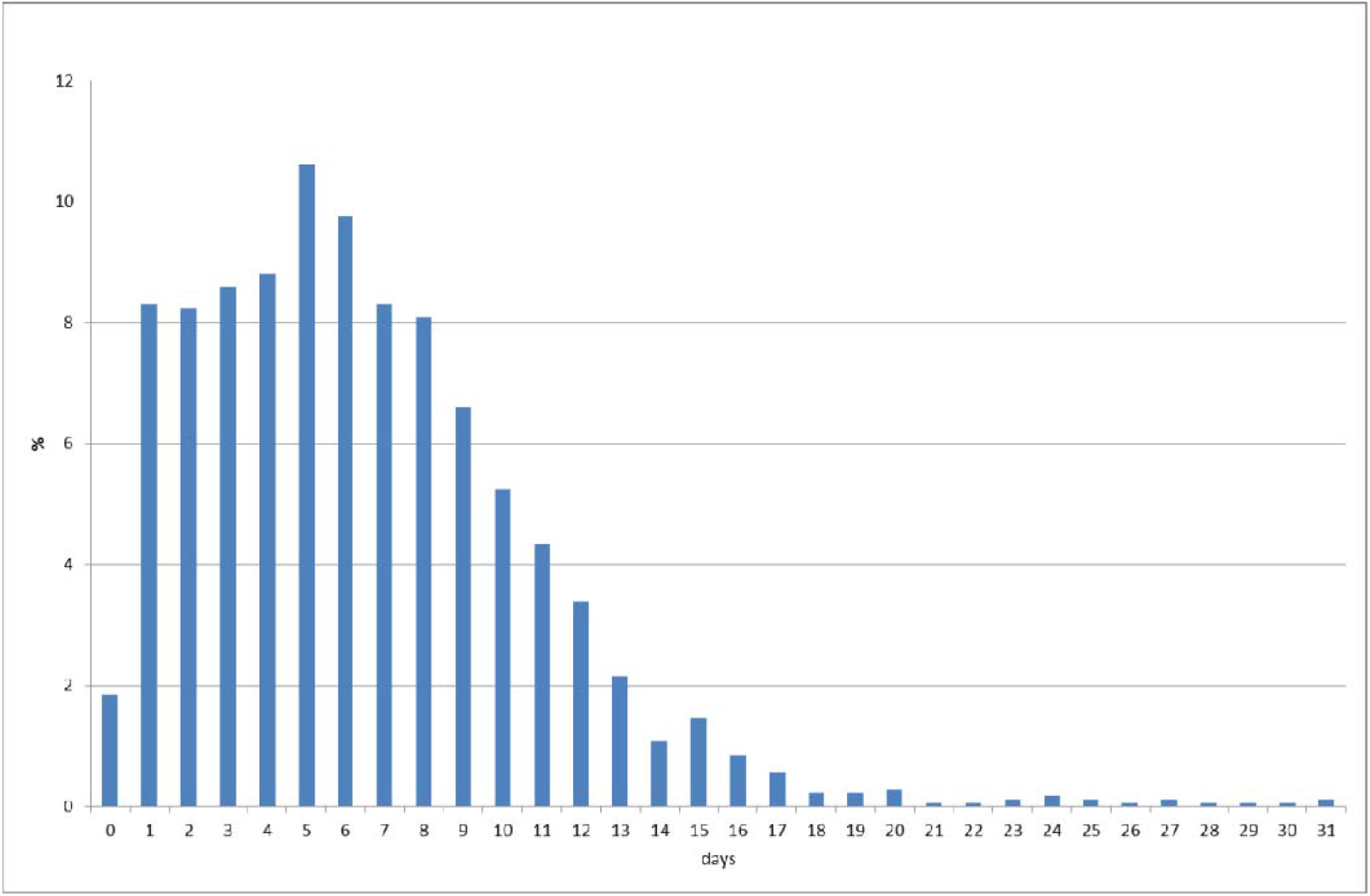
Empirical distribution of durations from onset to report by MHLW, Japan. (%) Note: Bars represent the probabilities of duration from onset to report based on data of 657 patients in Japan for whom onset dates were available. The horizontal axis shows the duration from exposure to onset in days. The vertical axis shows the probability of the incubation period. Data were obtained from MHLW, Japan.

**Fig. 3.**
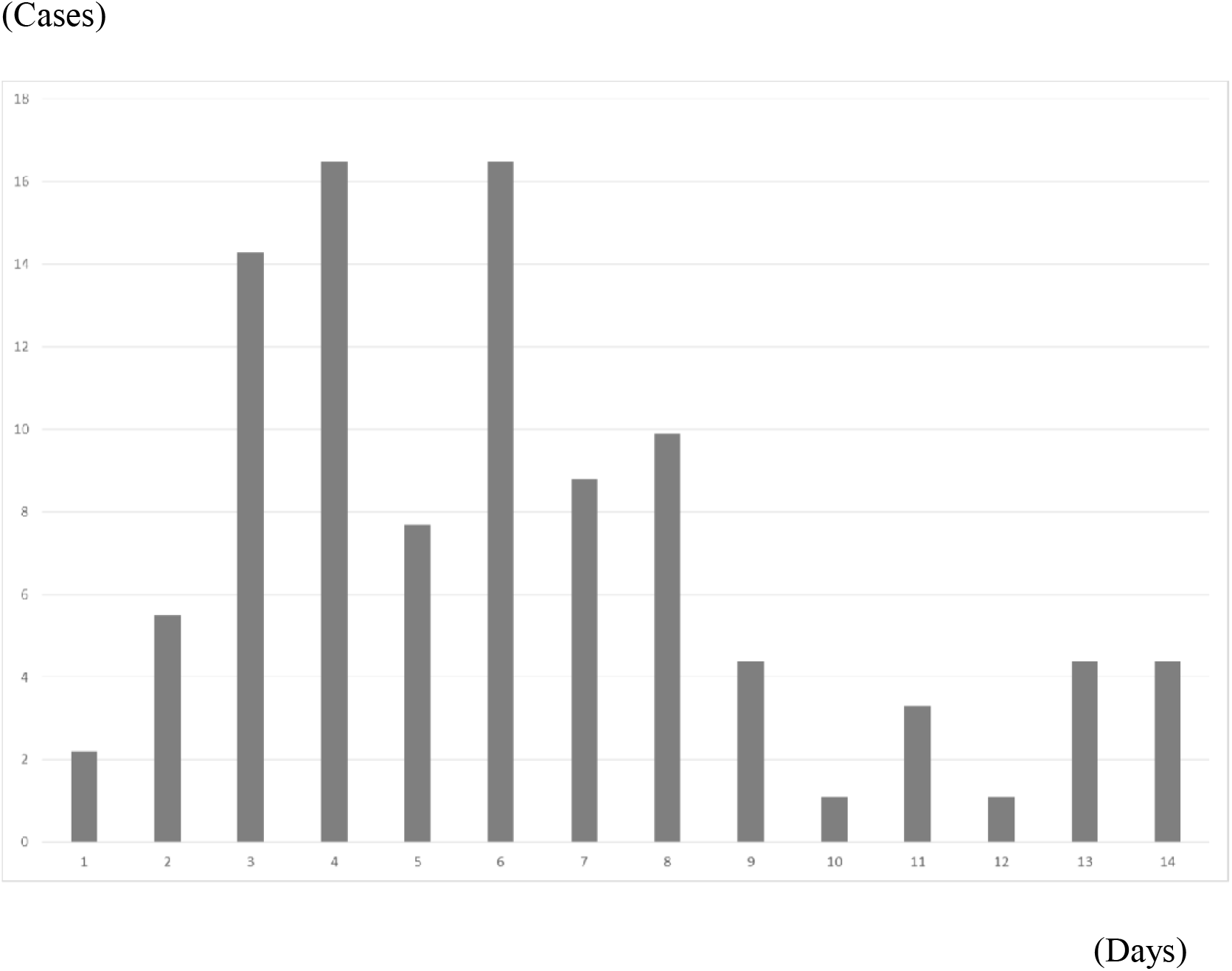
Empirical distribution of the incubation period published by MHLW, Japan. (%) Notes: Bars show the distribution of incubation periods for 125 cases for which the exposure date and onset date were published by MHLW, Japan. The horizontal axis was measured in duration from exposure to onset in days. The vertical axis was the probability of the incubation period. Patients for whom incubation was longer than 14 days are included in the bar shown for day 14.

The estimated reproduction number of the variant strain of SARS-CoV-2 was 1.799; its 95% CI was [1.642, 1.938]. The onset date of the first case in Japan was estimated as December 4 ([November 16, December 14]), 2020. However, the infectiousness of the original strain was estimated as 1.123 ([1.093, 1.166]). Figures 4 and 5 show the fitted line and its 95% CI of the number of laboratory-confirmed cases by the model for the two strains. Although stepwise data were difficult to mimic by the model, bands of CI fairly covered the observation data.

**Fig. 4.**
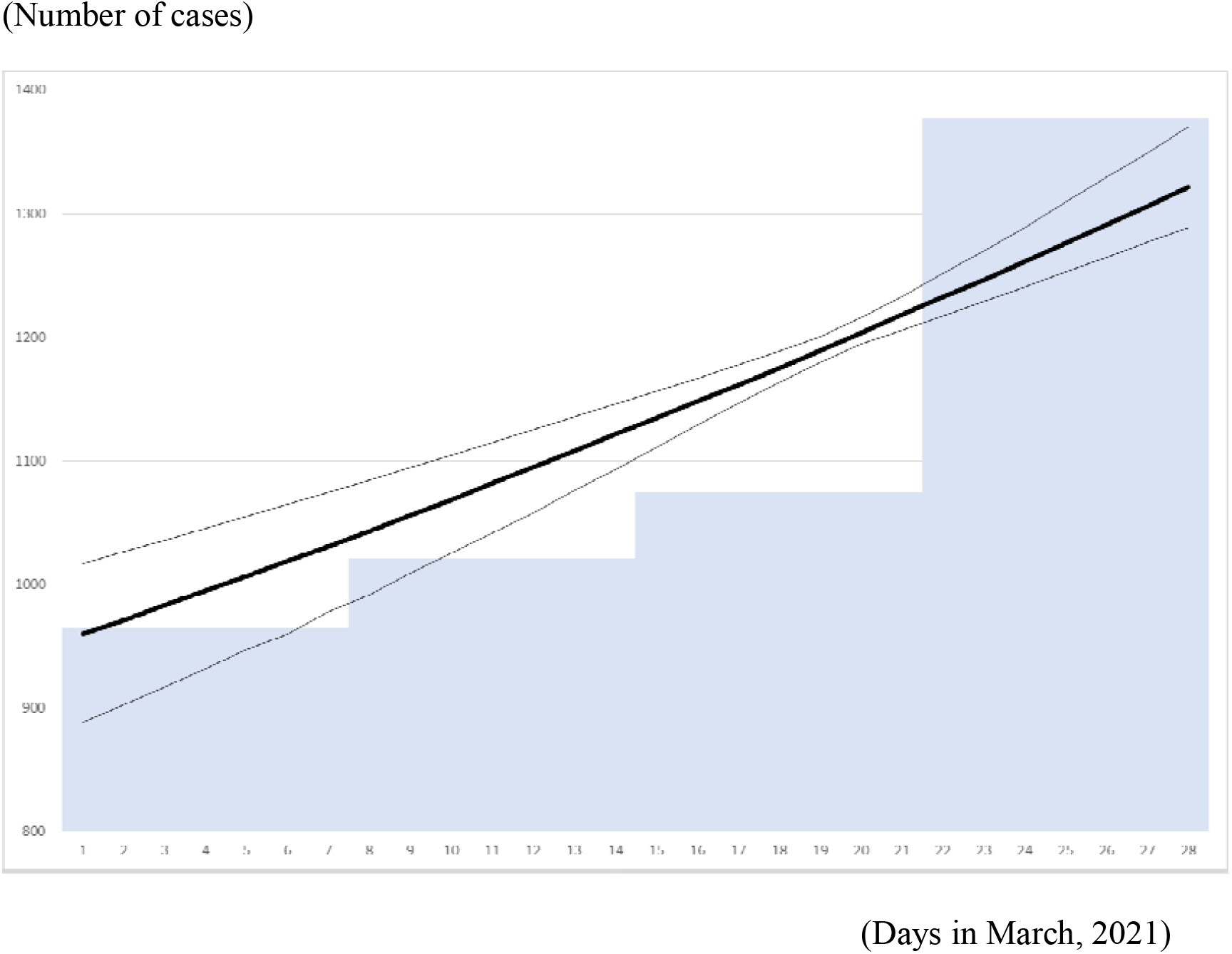
Observed data of the original strain and prediction with 95% CI Note; Bars represent the reported number of the test confirmed cases. The bold line represents its prediction by the model. Thin lines represent its 95% CI.

**Fig. 5.**
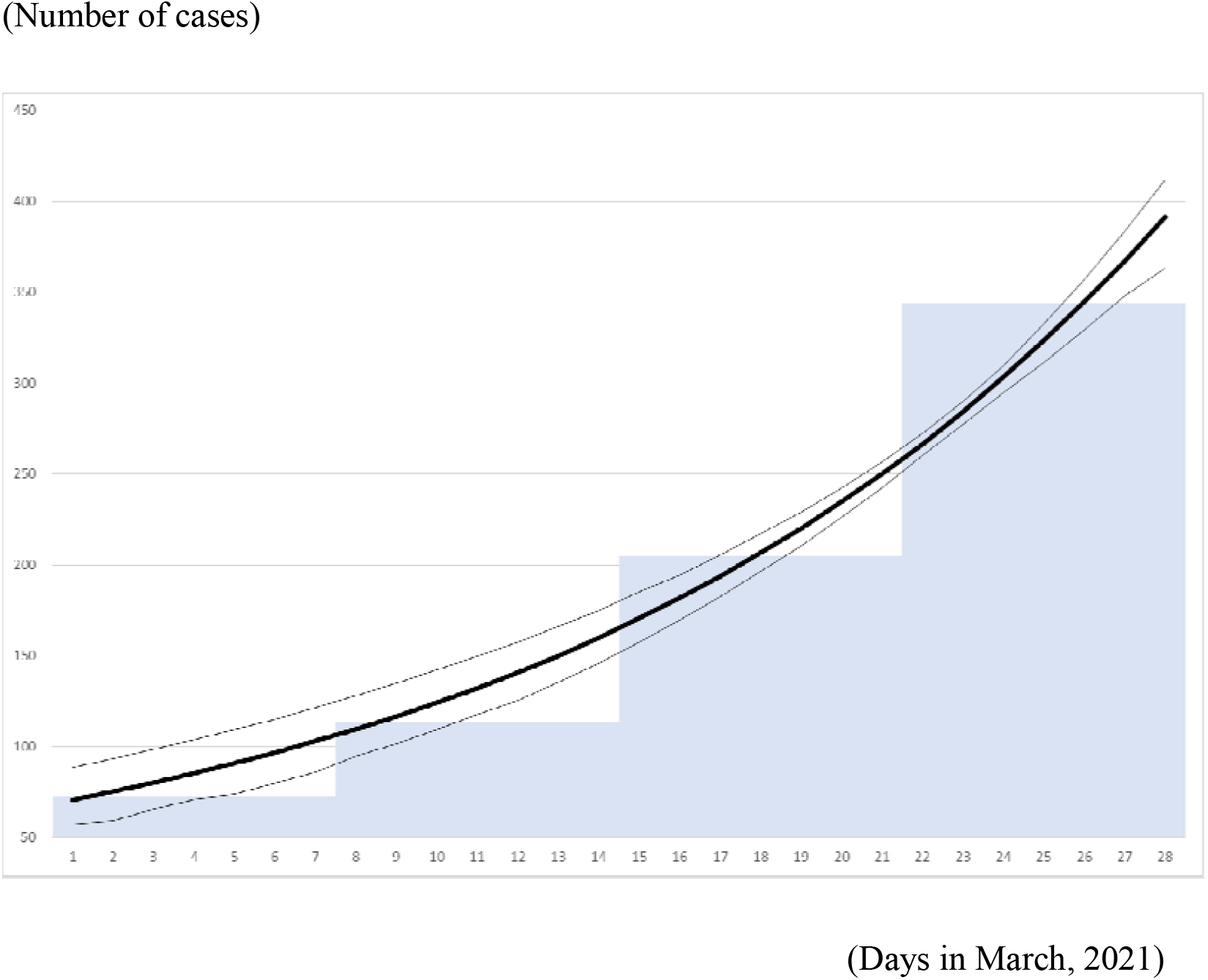
Observed data of the variant strain and its prediction with 95% CI. Note: Bars represent the reported numbers of the test confirmed cases. Bold line represents its prediction by the model. Thin lines represent 95% CI.

## Discussion

Similar differences of infectiousness were found for the variant strain and the original strain, as described for an earlier study [1]. The infectiousness increased by 0.684, indicating a 60% increase from the original to the variant. The rate of increase was slightly lower than that presented in an earlier report [1]. It might reflect the outbreak situation including vaccine coverage.

The vaccine coverage in March, 2021 in Japan was less than 1%. Therefore, it probably did not affect the outbreak at all. However, with increasing vaccine coverage in Japan, differences among strains are not expected to be affected if the vaccine effectiveness is equal for all strains. Alternatively, differences among strains will become increasingly evident if vaccine effectiveness against variant strains is less than the effectiveness against the original strain.

We estimated the onset date of the initial case of the community outbreak by a variant strain as December 4 ([November 16, December 14]), 2020. Actually, the first case of the variant strain was detected on January 6, 2021 as an imported case from UK. Therefore, we predicted the initial case of the variant strain as earlier than one month from the officially reported imported case. As one might expect, the earlier stage of the outbreak in the variant strain was not recognized in December, 2020 and January, 2021 because viral genomic sequencing, which can detect a variant strain, was not applied to the specimen from the community outbreak in Japan. Therefore, we cannot deny the possibility that a community outbreak by the variant strain occurred earlier than December, 2020, in Japan. The earliest case with the variant strain in the UK was reportedly identified on September 20, 2020 [1]. Therefore, no community outbreak was likely to have started in Japan two months later than in the UK.

The present study has some limitations. First, because of data limitations, some differences among variant strains were not considered. However, examination of those differences might be important for predicting which subtype of variant strain is dominant in the outbreak.

Secondly, the obtained result reflects information up to March, 2021. Changes in climate or mobility might have affected the results. In addition, as described above, increasing vaccine coverage might eventually affect results. Therefore, it is necessary to monitor differences of infectiousness among strains carefully and continuously.

## Conclusion

We demonstrated that infectiousness of the variant was greater by 0.684, or 60%, compared to the original strain of COVID-19. The findings might be comparable to those of a study conducted in the UK [1]. Nevertheless, careful and continuous monitoring of differences among strains is necessary.

The present study is based on the authors’ opinions. Its results do not reflect any stance or policy of their professionally affiliated bodies.

## Data Availability

Ministry of Health, Labour and Welfare, Implementation rate and positive rate of screening test for the variant strain (mechanical estimation) preliminary figures. (in Japanese)
World Health Organization. Coronavirus disease (COVID-19) Weekly Epidemiological Update and Weekly Operational Update

https://www.mhlw.go.jp/content/000766381.pdf

https://www.who.int/emergencies/diseases/novel-coronavirus-2019/situation-reports

## Acknowledgments

We acknowledge the great efforts of all staff at public health centers, medical institutions, and other facilities who are fighting the spread and destruction associated with COVID-19.

